# Estimated Effects of Amyloid Reduction on Cognitive Change: A Bayesian Update across a Range of Priors

**DOI:** 10.1101/2023.04.28.23289223

**Authors:** Sarah F. Ackley, Jingxuan Wang, Ruijia Chen, Melinda C. Power, Isabel Elaine Allen, M. Maria Glymour

## Abstract

**Introduction:** Results of the CLARITY-AD and GRADUATE I and II trials rekindled discussion on the impact of amyloid-targeting drugs. We use a Bayesian approach to quantify how a rational observer would have updated their prior beliefs based on new trial results.

**Methods:** We used publicly available data from the CLARITY-AD and GRADUATE I & II trials to estimate the effect of reducing amyloid on CDR-SB score. A range of prior positions were then updated according to Bayes Theorem using these estimates.

**Results:** After updating with new trial data, a wide range of starting positions resulted in credible intervals that did not include no effect of amyloid reduction on CDR-SB.

**Discussion:** For a range of starting beliefs and assuming veracity of underlying data, rational observers would conclude there is a small benefit of amyloid-reductions on cognition. This benefit must be weighed against opportunity cost and side effect risk.

## Introduction

The results of the CLARITY-AD trial rekindled discussion and debate on the value of reducing brain amyloid levels with amyloid-targeting drugs (1–4). Previous meta-analyses of randomized trials of amyloid-targeting drugs suggested the effects of amyloid-removal on cognitive outcomes were close to null (5–10), and these studies continue to be cited and referenced as evidence that an overall effect of this mechanism is likely to be negligible. Some have interpreted the small statistically significant effects reported in CLARITY-AD as unconvincing. While some cite concerns regarding publications bias and other design issues (11), others cite lack of clinical relevance of the reported effect size (12). Common counter arguments include that newer monoclonal antibody drugs targeting protofibrils instead of other amyloid species may confer benefits that earlier drug classes did not, as they can achieve greater reductions in the relevant species that is not necessarily captured by simple measure of overall change in amyloid (13–15) or because of nonlinear effects such that the cognitive benefits can only be achieved with the amyloid reductions produced by these newer medications (16).

Bayesian statistics, formalized from the early to mid-twentieth century (17–19), introduced the concept of a rational observer, which describes an individual who will constrain and update their beliefs according to the rules of probability (20). Rational observers may have radically different starting beliefs, but, as increasing evidence accumulates, rational observers ultimately converge on the truth. In such a model, beliefs described with a *prior* probability distribution are updated with new evidence in order to obtain a *posterior* probability distribution that describes updated beliefs (19).

Here, we consider how rational observers with very different beliefs about the effect of reducing amyloid on cognition, which may arise due to differences in beliefs about the pathogenesis of Alzheimer’s disease (2–4), publication bias (21), or trustworthiness of pharmaceutical companies (e.g., in reporting deaths (22)), would have responded to the recently announced results from trials of lecanemab (CLARITY-AD in September of 2022) and gantenerumab (GRADUATE I and II in December of 2022). The goal of this analysis is to systematically describe how beliefs should have been updated when combining recent evidence with a range of starting beliefs about amyloid reduction.

## Methods

Summary statistics from the CLARITY-AD trial and GRADUATE I & II trials were used to obtain estimates of CDR-SB (clinical dementia rating scale, sum of boxes score) change per 10 centiloid reduction. Since only aggregated data was available and presented inconsistently across these trials, we used previously developed instrumental variable (IV) methods to estimate these quantities (5,23). These IV methods were developed to use aggregated data to obtain a standardized effect of amyloid removal on cognitive change across trials with heterogeneous amyloid removal (24). CDR-SB scores are on an 18 point scale, reflecting that scores out of six were obtained from three reviewers. Increases on the CDR-SB scale are indicative of cognitive worsening. Summary statistics used to produce these estimates and associated standard errors are given in the Supplemental Methods (1,25).

We consider four initial positions for beliefs about the effect of reducing amyloid on cognition: a position based on results of previous meta-analyses (10,24), a position based on an assumption of significant publication bias that ascribes equal probability to benefit and harm, a position based on putative harm, and a position based solely on studies of monoclonal antibodies (24). Each position was given a normally distributed prior, with equal variances for all four positions. Table 1 describes the rationales for these priors and gives the means of each prior and symmetric intervals within which 95% of the probability mass of the prior distribution is located. This interval is a credible interval since we are explicitly ascribing belief to these priors. Additional priors with a range of means and variances were used in sensitivity analyses to validate the results.

**Table 1:** Description of prior positions and mean and 95% credible interval for the effect of a 10 centiloid reduction on CDR-SB change for each of the four main starting positions. All priors have the same variance.

| Position | Rationale | Prior Mean and 95% Credible Interval |
| --- | --- | --- |
| A. Meta-Analysis Informed | A prior meta-analysis gave a point estimate corresponding to a small benefit (based on Ackley 2021). We use the confidence interval from this meta-analysis as the basis for our prior. | -0.028<br>(-0.072, 0.015) |
| B. Publication Bias | Skeptics may believe prior meta-analyses were too optimistic, given the potential for publication bias and numerous trials registered in clinicaltrials.gov with no results ever reported. We adopt a prior with the same variance as in the meta-analysis-informed case, centered at no effect and ranging from small harm to small benefit. | 0.000<br>(-0.043, 0.043) |
| C. Putative Harm | Given concerns about side effects of amyloid-targeting therapies, some individuals may believe such therapies are harmful. We adopt a credible interval with the same variance as in the meta-analysis-informed case, with uncertainty ranging from very small to moderate sized harms. | 0.055<br>(0.012, 0.099) |
| D. Based on Monoclonal Antibody Drugs (MAB) | Some researchers prefer meta-analyses restricted to data from select trials of monoclonal antibody drugs, which indicate a larger benefit than the comprehensive meta-analysis previously considered. We adopt a posterior with the same variance as in the meta-analysis-informed case, with uncertainty ranging from very small to moderate sized benefits. | -0.055<br>(-0.099, -0.012) |

The four prior positions were updated based on Bayes Theorem using the IV estimates for the effect of a 10 centiloid reduction on the CDR-SB score from the CLARITY-AD trial and GRADUATE I & II trials. We used the fact that the normal distribution is a conjugate prior for a normally distributed likelihood function with a fixed variance in order to have a closed-form solution for the posterior distribution. This gives an analytic solution for the posterior distribution, which is also normal (26). The likelihood was considered to be normally distributed with mean given by the effect per 10 centiloids estimated from the trials and variance. Since the *t*-distribution does not have a conjugate prior (27), the variance was assumed to be precisely known and given by a variance obtained from IV estimates. This is analogous to using *z* vs *t* statistics in frequentist statistics, and, given the size of the trials, can be considered a very good approximation. The prior position was updated with both the CLARITY-AD trial and the GRADUATE I & II trials together, as well as with CLARITY-AD trial and the GRADUATE I and II trials individually.

As sensitivity analyses, we consider 33 additional starting priors, ranging from effects of -0.1 to 0.1 CDR-SB points per 10 centiloid reduction. We also evaluated alternate variances–both narrower and wider by a factor of two than that of the main analysis–for this range of prior means. These priors are specified in Supplemental Table S1.

Since this study used only publicly available, aggregated summary statistics, this study is not human subjects research. All analysis was performed using R version 4.2.1.

## Results

The IV estimates for the effect of a 10 centiloid reduction on CDR-SB scores are -0.076 (standard error, 0.019) for CLARITY-AD (lecanemab) and -0.043 (standard error, 0.021) for GRADUATE I & II (gantenerumab). Means and associated standard errors were used to define the likelihood used to update the starting priors. Posterior means and associated credible intervals for updates of the four main priors with estimates from CLARITY-AD and GRADUATE I & II estimates are shown in Table 2. Prior and posterior distributions are shown in Figure 1. After updating with only the CLARITY-AD estimates, the mean of the posteriors varied from a benefit of 0.020 to 0.067 CDR-SB points per 10 centiloid reduction, with only the 95% credible interval for the putative harm position including no effect. After updating with only the GRADUATE I & II estimate, the mean of the posteriors varied from a harm of 0.003 to a benefit of 0.049, with 95% credible intervals for the publication bias and putative harm positions including no effect. After updating with both CLARITY-AD and GRADUATE I & II data the mean of the posteriors varied from a benefit of 0.027 to 0.059, and no 95% credible intervals contained no effect. Table and Figure S1 give the credible intervals posterior means and associated credible intervals for updates of additional priors.

**Table 2:** Prior and posterior means and credible intervals for the effect of a 10 centiloid reduction on CDR-SB for each of the four main starting positions. Posterior columns give means and credible intervals for posteriors updated with just the CLARITY-AD estimates, posteroirs updated with just the GRAUDATE I & II estimates, and posteriors updated with both the CLARITY-AD and GRADUATE I & II estimates. All prior and posterior distributions are normally distributed and the interval is constructed to be symmetric about the mean and contain 95% of the probability mass. MAB: monoclonal antibodies.

| Prior Description | Prior | Posterior CLARITY-AD Only | Posterior GRAUDATE I & II Only | Posterior CLARITY-AD and GRAUDATE I & II |
| --- | --- | --- | --- | --- |
| A. Meta-Analysis Informed | -0.028<br>(-0.072, 0.015) | -0.056<br>(-0.084, -0.027) | -0.036<br>(-0.066, -0.006) | -0.051<br>(-0.075, -0.028) |
| B. Publication Bias | 0.000<br>(-0.043, 0.043) | -0.044<br>(-0.072, -0.015) | -0.023<br>(-0.052, 0.007) | -0.043<br>(-0.067, -0.020) |
| C. Putative Harm | 0.055<br>(0.012, 0.099) | -0.020<br>(-0.048, 0.008) | 0.003<br>(-0.027, 0.033) | -0.027<br>(-0.051, -0.004) |
| D. Based on MAB | -0.055<br>(-0.099, -0.012) | -0.067<br>(-0.096, -0.039) | -0.049<br>(-0.078, -0.019) | -0.059<br>(-0.083, -0.036) |

**Figure 1:**
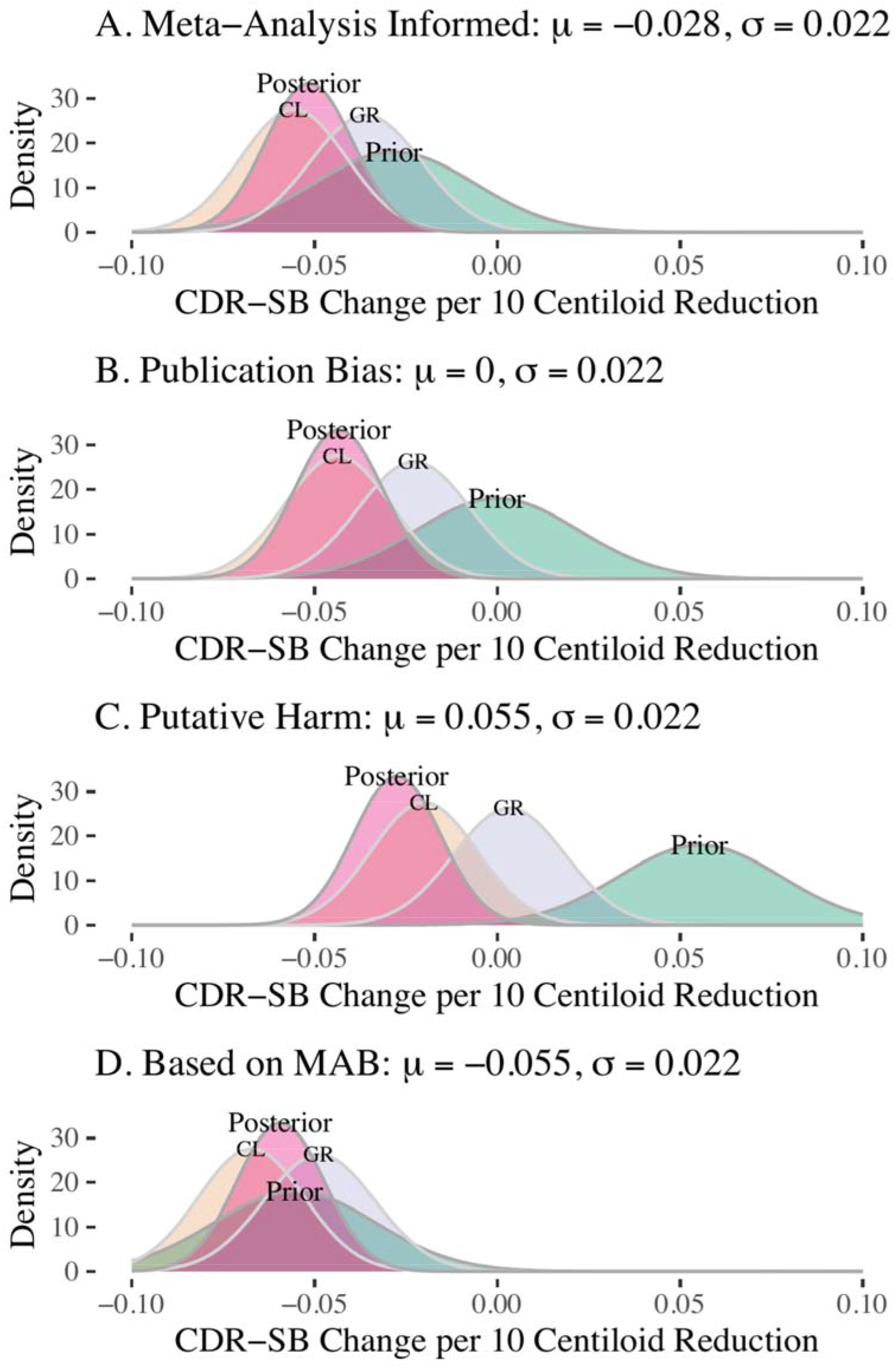
Prior intervals and posterior distributions for the effect of a 10 centiloid reduction on CDR-SB for the four main starting positions. Panels A-D correspond to the four starting positions outlined in Tables 1 and 2. Each panel shows the prior, a posterior updated with just the CLARITY-AD estimates (labled “CL”), a posteroir updated with just the GRAUDATE I & II estimates (labeled “GR”), and a final posterior (labeled “Posterior”) that updates with both the CLARITY-AD and GRADUATE I & II estimates. Mean () and standard deviation () of the prior are given in the panel label. MAB: monoclonal antibodies.

## Discussion

After updating with CLARITY-AD and GRADUATE findings, all four starting positions resulted in credible intervals that did not include no effect of amyloid reduction on CDR-SB change. Although the most divergent starting positions differed by 0.110, means of the posteriors differed by only 0.032 CDR-SB points per 10 centiloid reduction. Therefore, for a wide range of starting beliefs, rational observers with those beliefs should conclude–if drawing inferences only from the available trial evidence–that within the timeframe of the trials there is a small benefit of amyloid-reductions due to amyloid targeting drugs on cognition. However, determining whether an individual amyloid-targeting drug has value requires consideration of not only the average effect, but whether the drug produces meaningful changes for an individual, the likelihood and severity of side effects, and the opportunity cost to the individual and to the healthcare system of investing resources in this drug rather than other aspects of treatment and care.

Many have argued that monoclonal antibody drugs or drugs that effectively target protofibrils are more effective than older small-molecule drugs to explain why prior trials failed. However, these arguments are no longer necessary to justify beliefs that amyloid targeting drugs are effective. This is because, with the addition of current evidence, pooling across all drugs for which there is data available indicates overall benefit. That is, for a wide range of starting beliefs, including those that equally weight past and current evidence, updated beliefs should assign low probabilities to no effect or cognitive harm of amyloid reduction. Further, the new posterior point estimate and credible interval are consistent with previous meta-analysis results showing small benefit across trials with confidence intervals that included the null because, without the new trial data, estimates were less precise (5).

Rational observers should also conclude that large benefits–i.e., those exceeding 0.1 CDR-SB points per 10-centiloid reduction–are also implausible. The most optimistic starting beliefs, based on the results of the EMERGE and ENGAGE trials of aducanumab, also preclude large benefits. For context, individuals with a diagnosis of symptomatic Alzheimer’s disease decline by an average of one to two CDR-SB points per year, depending on severity (28). In addition, although the GRADUATE I & II trials did not meet their primary endpoint (29,30), the point estimates were similar to the meta-analyzed effect estimate and narrowed the credible intervals around that estimate to give a credible interval that did not include no effect for the meta-analysis informed starting position. That is, while gantenerumab’s effects in GRADUATE I & II appear to be smaller than those of CLARITY-AD, in the context of prior evidence, they do support a small overall benefit of amyloid-reducing drugs.

Our analysis has important limitations. Priors outside the range of those we considered or violations of other necessary assumptions may imply that observers may conclude that amyloid-targeting drugs confer no benefit, despite the CLARITY-AD and GRADUATE results. For example, some observers may have started with a prior that indicated large harm, exceeding that of the putative harm starting position, despite inconsistency for this prior with meta-analyses to date and evidence against publication or reporting bias hiding a significant, harmful effect (including the fact that significant cognitive harm was reported for BACE inhibitors (31), and the fact that, trials of drugs causing significant harm or significant benefit are most likely to be reported on–at least in some capacity). While we assume veracity of reported data, skeptics may believe that the CLARITY-AD and GRADUATE trials were poorly designed or had significant flaws leading to biased estimates. This might include loss of blinding, broken randomization, differential loss-to-follow-up or mortality, or frank malfeasance and data fabrication (1,11). It is beyond the scope of this article to assess whether data quality concerns are plausible or have been adequately addressed (e.g., sensitivity analyses in supplemental information of (1)). While not currently achieved on this topic, we encourage trial investigators to be open to reporting that extends beyond accurately presenting methods and results. Current skepticism is fueled by financial conflicts of interest, lack of long-term followup, and lack of data sharing (21). Providing extra evidence (i.e., data sharing) and consideration would help the field come to consensus.

Individuals have a range of beliefs on the effect of amyloid removal on cognition. The paper represents an attempt to ascertain whether consensus on whether amyloid removal reduces cognitive decline is reasonable given the diversity of starting opinions and based on the available trial data to date. Our results would indicate that amyloid removal appears to confer small benefit based on available evidence, but do not pertain to the value of any particular drug. Debates will certainly continue regarding whether these drugs should be approved or reimbursed and in which populations, as such decisions require not only evidence of an effect, but the clinical relevance of an effect, safety concerns, and other factors. Moreover, individual decisions about whether to take an approved drug will additionally involve consideration of the associated opportunity costs. Specifically, questions remain regarding safety and concurrent anticoagulant usage (22), generalizability of findings to Black and other populations with increased vascular risk (32), costs of the drug and infrastructure required MRI safety monitoring (33), and long-term outcomes (34,35).

The results of this study are not surprising in the context of prior frequentist meta-analyses. In Ackley et al., the confidence interval for the overall effect of amyloid reduction on cognitive change in 2021 included both small effects and no effect of amyloid reduction on cognitive change (0.05, 95% CI: (-0.032, 0.13) MMSE points per centiloid). However, that meta-analysis included a calculation that suggested one compelling trial could shift the overall estimate to statistical significance. A meta-analysis by Pang et al. replicating Ackley et al. included data from the phase II PRIME trial of aducanumab, one of the most optimistic trials of the benefits of amyloid removal to date (cognitive change per SUVR change), and obtained an estimate that indicated overall benefit (23). Other analyses did not estimate a cognitive benefit per amyloid change, but indicated either null effects or small benefits were plausible (6–10). None of these meta-analyses include the most recent data from CLARITY-AD (1) or GRADUATE I & II (unpublished).

Some researchers argue based on qualitative assessment of the trials (36) and animal models that amyloid removal has nonlinear effects on cognition, with benefits only becoming apparent once a large fraction of amyloid has been removed (16). Reporting a per 10 centiloid decrease could be viewed as an inaccurate linearity assumption. However, we note that CLARITY-AD and GRADUATE I & II removed a similar amount of amyloid and thus our estimates pertain to the specific ranges of amyloid in these trials. Linearity assumptions would only imply a potential for the irrationality of a meta-analysis-informed and more pessimistic starting positions. This position was informed by 15 trials, including trials for which drug treatment removed very little amyloid. However, in this case this would represent a conservative bias. That is, this would make it less likely that we would ascribe low probability to amyloid reduction either having no effect or being harmful. However, for this and more pessimistic positions, less than a 2.5% probability was assigned to the possibility that amyloid reduction had either no effect or was harmful. So even under the potentially conservative biases of the first three starting positions, we concluded benefits of amyloid removal were likely.

This quantitative approach allows for a determination of rational positions given a range of prior starting beliefs. Even for rational observers who believe amyloid reduction is more likely harmful than not, the recent trials provide evidence to shift beliefs from likely harm to likely benefit. Current debates should focus on upstream issues such as data and analysis quality and downstream issues such as whether minimum clinically meaningful differences have been achieved, safety, infrastructure, and generalizability of results. Based on available evidence, it seems reasonable to believe that amyloid reduction is helpful but provides at most small cognitive benefit on the timescale of these trials.

## Data Availability

All data produced in the present work are contained in the manuscript.

https://www.alzforum.org/news/conference-coverage/gantenerumab-mystery-how-did-it-lose-potency-phase-3

https://www.nejm.org/doi/full/10.1056/NEJMoa2212948

## Acknowledgements/Conflicts/Funding Sources

This work was supported by the National Institute of Aging (NIA) under award numbers R01AG057869 (MMG, MCP), K99AG073454 (SFA), and K00AG068431 (RC).

## Supplemental Methods

Data from the CLARITY-AD trial and GRADUATE I and II trials used to estimate the effect of a 10 centiloid reduction in amyloid on the CDR-SB score and associated standard errors.

- CLARITY-AD trial (1):
  - CRD-SB change: -0.45 with the 95% CI: (-0.67, 0-.23) used to generate a standard error using the quantile function of the normal distribution.
  - Centiloid change: -59.12 with the 95% CI: (-62.64, -55.60) used to generate a standard error using the quantile function of the normal distribution.
- GRADUATE I and II trials (25):
  - Overall CDR-SB change: -0.26 with p=0.04 used to generate a standard error.
  - Centiloid change placebo groups: 8.5 and 8.7.
  - Centiloid change treatment groups: -46.8 and -57.6.
  - The overall centiloid difference between groups was assumed to be the unweighted average of centiloid differences in GRADUATE I and II. I.e., - ((46.8+8.5)/2 + (57.6+8.7)/2).
  - The standard deviation in centiloid change was assumed to be the same as in the CLARITY-AD trial. This was used to generate a standard error using n=1795 (CLARITY-AD) and n=1965 (GRADUATE I and II). That is, the gantenerumab standard error in amyloid change was assumed to be lecanemab standard error times the square root of 1965/1795.

## Supplemental Tables and Figures

**Table S1:**
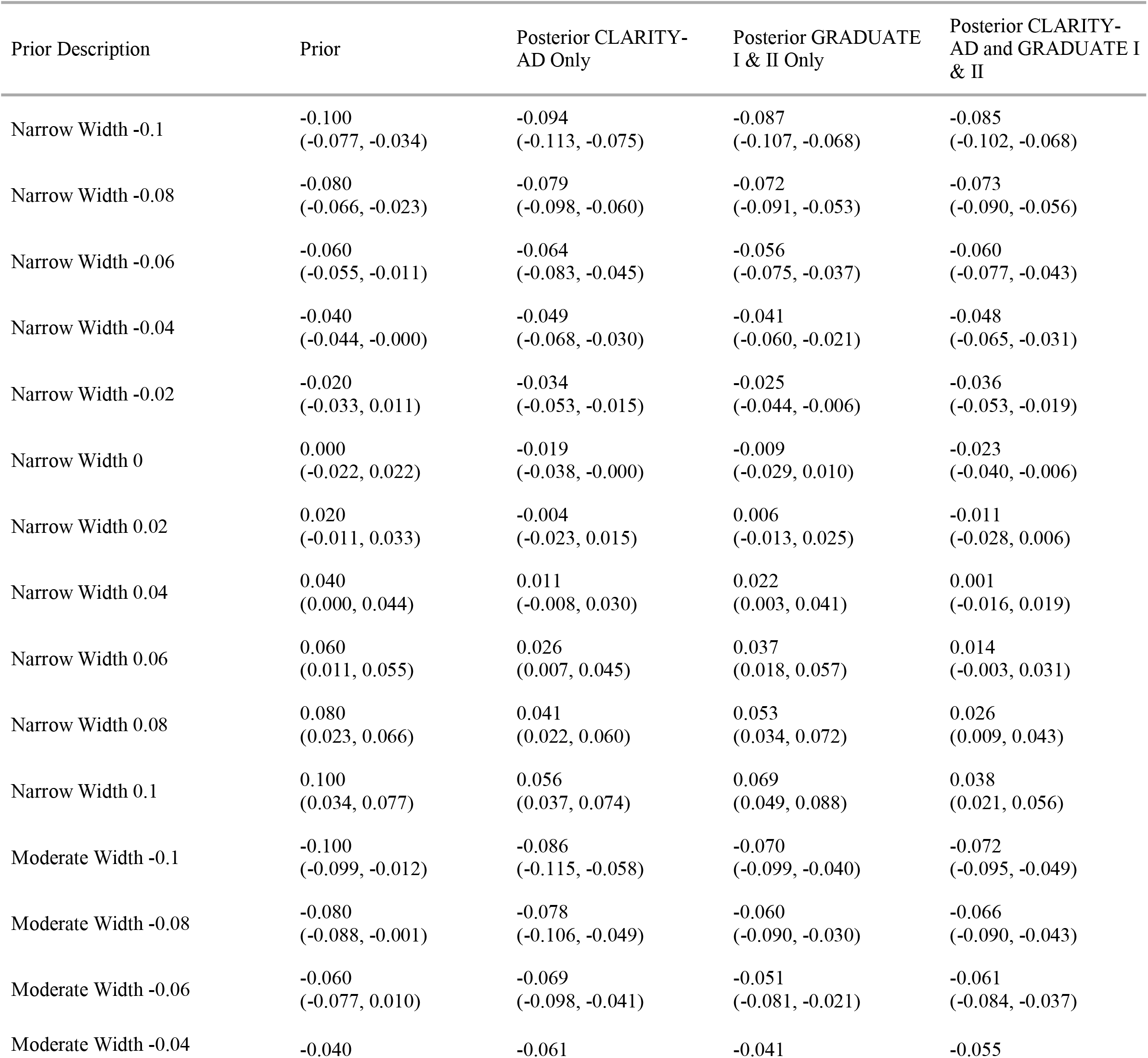
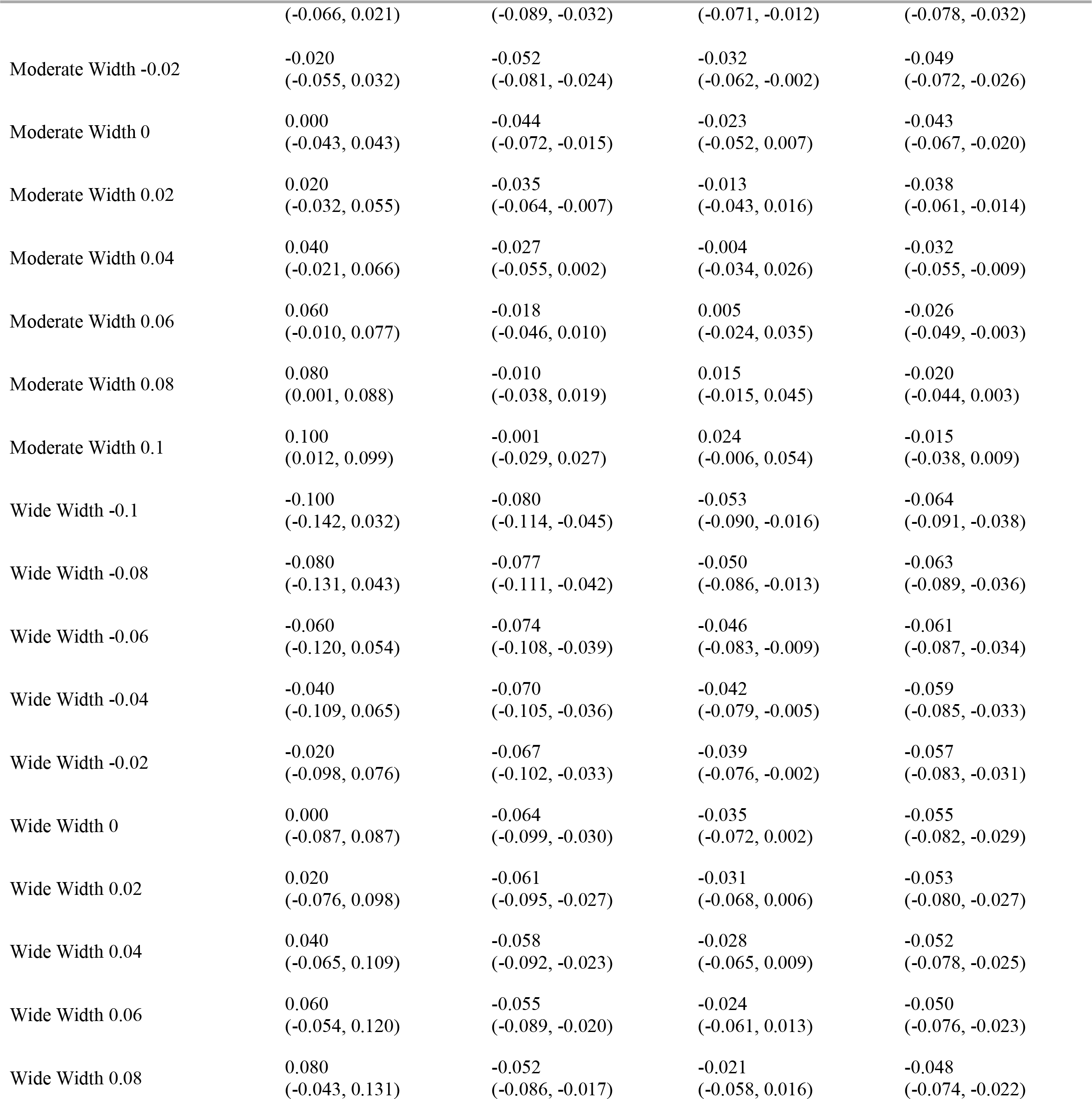
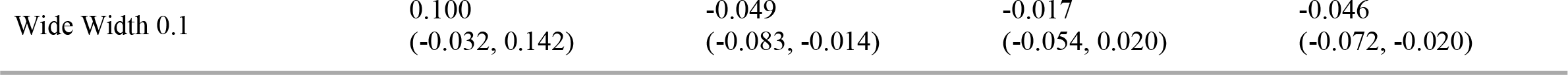
Prior and posterior means and credible intervals for the effect of a 10 centiloid reduction on CDR-SB for narrow, moderate, and wide starting prior distributions with mean given in the description. All prior and posterior distributions are normally distributed and the interval is constructed to be symmetric about the mean and contain 95% of the probability mass.

| Prior Description | Prior | Posterior CLARITY-AD Only | Posterior GRADUATE I & II Only | Posterior CLARITY-AD and GRADUATE I & II |
| --- | --- | --- | --- | --- |
| Narrow Width -0.1 | -0.100<br>(-0.077, -0.034) | -0.094<br>(-0.113, -0.075) | -0.087<br>(-0.107, -0.068) | -0.085<br>(-0.102, -0.068) |
| Narrow Width -0.08 | -0.080<br>(-0.066, -0.023) | -0.079<br>(-0.098, -0.060) | -0.072<br>(-0.091, -0.053) | -0.073<br>(-0.090, -0.056) |
| Narrow Width -0.06 | -0.060<br>(-0.055, -0.011) | -0.064<br>(-0.083, -0.045) | -0.056<br>(-0.075, -0.037) | -0.060<br>(-0.077, -0.043) |
| Narrow Width -0.04 | -0.040<br>(-0.044, -0.000) | -0.049<br>(-0.068, -0.030) | -0.041<br>(-0.060, -0.021) | -0.048<br>(-0.065, -0.031) |
| Narrow Width -0.02 | -0.020<br>(-0.033, 0.011) | -0.034<br>(-0.053, -0.015) | -0.025<br>(-0.044, -0.006) | -0.036<br>(-0.053, -0.019) |
| Narrow Width 0 | 0.000<br>(-0.022, 0.022) | -0.019<br>(-0.038, -0.000) | -0.009<br>(-0.029, 0.010) | -0.023<br>(-0.040, -0.006) |
| Narrow Width 0.02 | 0.020<br>(-0.011, 0.033) | -0.004<br>(-0.023, 0.015) | 0.006<br>(-0.013, 0.025) | -0.011<br>(-0.028, 0.006) |
| Narrow Width 0.04 | 0.040<br>(0.000, 0.044) | 0.011<br>(-0.008, 0.030) | 0.022<br>(0.003, 0.041) | 0.001<br>(-0.016, 0.019) |
| Narrow Width 0.06 | 0.060<br>(0.011, 0.055) | 0.026<br>(0.007, 0.045) | 0.037<br>(0.018, 0.057) | 0.014<br>(-0.003, 0.031) |
| Narrow Width 0.08 | 0.080<br>(0.023, 0.066) | 0.041<br>(0.022, 0.060) | 0.053<br>(0.034, 0.072) | 0.026<br>(0.009, 0.043) |
| Narrow Width 0.1 | 0.100<br>(0.034, 0.077) | 0.056<br>(0.037, 0.074) | 0.069<br>(0.049, 0.088) | 0.038<br>(0.021, 0.056) |
| Moderate Width -0.1 | -0.100<br>(-0.099, -0.012) | -0.086<br>(-0.115, -0.058) | -0.070<br>(-0.099, -0.040) | -0.072<br>(-0.095, -0.049) |
| Moderate Width -0.08 | -0.080<br>(-0.088, -0.001) | -0.078<br>(-0.106, -0.049) | -0.060<br>(-0.090, -0.030) | -0.066<br>(-0.090, -0.043) |
| Moderate Width -0.06 | -0.060<br>(-0.077, 0.010) | -0.069<br>(-0.098, -0.041) | -0.051<br>(-0.081, -0.021) | -0.061<br>(-0.084, -0.037) |
| Moderate Width -0.04 | -0.040 | -0.061 | -0.041 | -0.055 |

| Prior Description | Prior | Posterior CLARITY-<br>AD Only | Posterior GRADUATE<br>I & II Only | Posterior CLARITY-<br>AD and GRADUATE I<br>& II |
| --- | --- | --- | --- | --- |
|  | (-0.066, 0.021) | (-0.089, -0.032) | (-0.071, -0.012) | (-0.078, -0.032) |
| Moderate Width -0.02 | -0.020<br>(-0.055, 0.032) | -0.052<br>(-0.081, -0.024) | -0.032<br>(-0.062, -0.002) | -0.049<br>(-0.072, -0.026) |
| Moderate Width 0 | 0.000<br>(-0.043, 0.043) | -0.044<br>(-0.072, -0.015) | -0.023<br>(-0.052, 0.007) | -0.043<br>(-0.067, -0.020) |
| Moderate Width 0.02 | 0.020<br>(-0.032, 0.055) | -0.035<br>(-0.064, -0.007) | -0.013<br>(-0.043, 0.016) | -0.038<br>(-0.061, -0.014) |
| Moderate Width 0.04 | 0.040<br>(-0.021, 0.066) | -0.027<br>(-0.055, 0.002) | -0.004<br>(-0.034, 0.026) | -0.032<br>(-0.055, -0.009) |
| Moderate Width 0.06 | 0.060<br>(-0.010, 0.077) | -0.018<br>(-0.046, 0.010) | 0.005<br>(-0.024, 0.035) | -0.026<br>(-0.049, -0.003) |
| Moderate Width 0.08 | 0.080<br>(0.001, 0.088) | -0.010<br>(-0.038, 0.019) | 0.015<br>(-0.015, 0.045) | -0.020<br>(-0.044, 0.003) |
| Moderate Width 0.1 | 0.100<br>(0.012, 0.099) | -0.001<br>(-0.029, 0.027) | 0.024<br>(-0.006, 0.054) | -0.015<br>(-0.038, 0.009) |
| Wide Width -0.1 | -0.100<br>(-0.142, 0.032) | -0.080<br>(-0.114, -0.045) | -0.053<br>(-0.090, -0.016) | -0.064<br>(-0.091, -0.038) |
| Wide Width -0.08 | -0.080<br>(-0.131, 0.043) | -0.077<br>(-0.111, -0.042) | -0.050<br>(-0.086, -0.013) | -0.063<br>(-0.089, -0.036) |
| Wide Width -0.06 | -0.060<br>(-0.120, 0.054) | -0.074<br>(-0.108, -0.039) | -0.046<br>(-0.083, -0.009) | -0.061<br>(-0.087, -0.034) |
| Wide Width -0.04 | -0.040<br>(-0.109, 0.065) | -0.070<br>(-0.105, -0.036) | -0.042<br>(-0.079, -0.005) | -0.059<br>(-0.085, -0.033) |
| Wide Width -0.02 | -0.020<br>(-0.098, 0.076) | -0.067<br>(-0.102, -0.033) | -0.039<br>(-0.076, -0.002) | -0.057<br>(-0.083, -0.031) |
| Wide Width 0 | 0.000<br>(-0.087, 0.087) | -0.064<br>(-0.099, -0.030) | -0.035<br>(-0.072, 0.002) | -0.055<br>(-0.082, -0.029) |
| Wide Width 0.02 | 0.020<br>(-0.076, 0.098) | -0.061<br>(-0.095, -0.027) | -0.031<br>(-0.068, 0.006) | -0.053<br>(-0.080, -0.027) |
| Wide Width 0.04 | 0.040<br>(-0.065, 0.109) | -0.058<br>(-0.092, -0.023) | -0.028<br>(-0.065, 0.009) | -0.052<br>(-0.078, -0.025) |
| Wide Width 0.06 | 0.060<br>(-0.054, 0.120) | -0.055<br>(-0.089, -0.020) | -0.024<br>(-0.061, 0.013) | -0.050<br>(-0.076, -0.023) |
| Wide Width 0.08 | 0.080<br>(-0.043, 0.131) | -0.052<br>(-0.086, -0.017) | -0.021<br>(-0.058, 0.016) | -0.048<br>(-0.074, -0.022) |
| Wide Width 0.1 | 0.100<br>(-0.032, 0.142) | -0.049<br>(-0.083, -0.014) | -0.017<br>(-0.054, 0.020) | -0.046<br>(-0.072, -0.020) |

**Figure S1:**
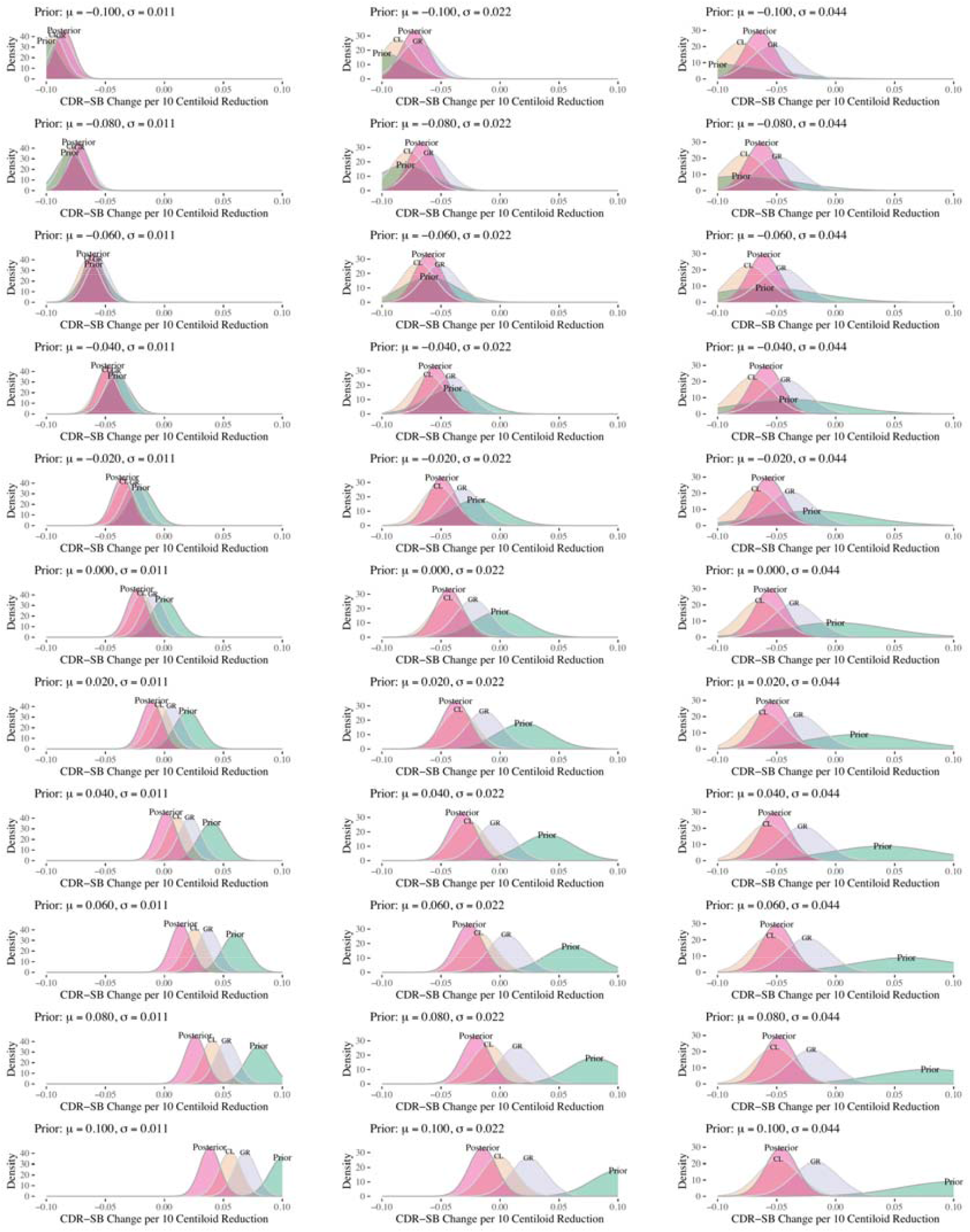
Prior and posterior distributions for the effect a 10 centiloid reduction on CDR-SB change for narrow, moderate, and wide starting prior distributions. Mean () and standard deviation () are given in the panel labels.

